# Ultrasound vs. MRI Assessment of Rotator Cuff Tear Severity: A Systematic Review and Meta-Analysis of Supraspinatus Tears

**DOI:** 10.64898/2026.09.12.26362920

**Authors:** Kareena Kassam, Shrimanti Ghosh, Stephanie Wichuk, Tina Qian, Jessica Knight, Cassandra Gallant, Steel McDonald, Jacob L. Jaremko, Abhilash R. Hareendranathan

## Abstract

**Objective:** Rotator cuff tears involving the supraspinatus tendon are a common cause of shoulder pain. Ultrasound (US) is accessible, dynamic, and lower cost than MRI, but diagnostic performance may vary by tear severity, operator expertise. This systematic review and meta-analysis compared the US with MRI for supraspinatus tear detection.

**Methods:** MEDLINE, Embase, Scopus, and CINAHL Plus were searched for studies published from January 2015 to June 2026. Study quality was assessed using QUADAS-2. Diagnostic outcomes included sensitivity, specificity, accuracy, PPV, NPV. Meta-analyses were performed separately for any, partial-thickness, and full-thickness tears when complete 2 × 2 data were available.

**Results:** 23 studies comprising 1,452 participants were included. US demonstrated stronger and more consistent performance for full-thickness than partial-thickness tears. Mean sensitivity, specificity, and accuracy were 88.5%, 95.2%, and 93.4%, respectively, for full-thickness tears, compared with 71.5%, 91.5%, and 79.2% for partial-thickness tears. HSROC analysis showed pooled sensitivity and specificity of 96% and 76% for any tear, 94% and 80% for partial-thickness tears, and 96% and 98% for full-thickness tears. Agreement measures also generally favored full-thickness tears. Operator expertise, study quality varied, and only one study evaluated a handheld ultrasound system.

**Conclusion:** Ultrasound demonstrates strong diagnostic performance for supraspinatus tears, particularly full-thickness tears, while partial-thickness tears remain more challenging. Given the high observed sensitivity, well-performed US may serve as a practical first-line modality to rule out full-thickness tears, with MRI remaining complementary for equivocal partial-thickness tears, associated pathology, and preoperative assessment.

## 1 Introduction

The shoulder is a highly mobile and complex joint whose function depends largely on the rotator cuff, comprising the supraspinatus, infraspinatus, teres minor, and subscapularis muscles and tendons [1, 2]. Rotator cuff tears (RCTs) are a common cause of shoulder pain and dysfunction in adults, accounting for 30–70% of cases [3]. The supraspinatus is most frequently affected because of its anatomical position and repetitive mechanical loading [3–5]. Tears may result from acute trauma or, more commonly, age-related degeneration, overuse, or impingement [1].

Athletes involved in overhead sports such as baseball, tennis, and swimming are particularly at risk of partial- and full-thickness tears [6, 7], which can impair performance and delay return to sport [6, 7]. Common symptoms include pain, weakness, limited motion, and functional impairment [8]. Although physical examination maneuvers such as the empty can test [9], drop arm test [10], and Neer sign [11] may suggest rotator cuff pathology, imaging is often required to confirm the diagnosis, determine tear severity, and guide management [12].

Distinguishing partial-from full-thickness tears is clinically important because tear severity influences management. Partial-thickness tears are often managed conservatively, whereas full-thickness tears may require surgical repair depending on symptoms and tear characteristics [13]. Although arthroscopy is considered the definitive reference standard for confirming rotator cuff tears, it is invasive and generally reserved for operative management. **MRI therefore serves as the principal non-invasive reference imaging modality for preoperative assessment**, providing detailed visualization of tear extent, tendon retraction, muscle atrophy, and associated pathology [14, 15]. Earlier reviews used arthroscopy as the definitive reference, supporting MRI as a suitable non-invasive comparator for US [16]. However, MRI is costly, may involve long waiting times, and may be unsuitable or less accessible for some patients [15, 17, 18].

Ultrasound (US) provides accessible, low-cost, real-time musculoskeletal (MSK) imaging [19], but remains operator-dependent and may be limited by obesity and reduced visualization of intra-articular structures [20, 21]. Despite these limitations, ultrasound remains more affordable and accessible than MRI, particularly with the increasing availability of handheld and point-of-care systems [22–24]. POCUS may further expand access to rapid bedside MSK assessment, although evidence for rotator cuff evaluation remains limited [25, 26]. Together, these advantages support ultrasound as a practical first-line imaging option for suspected rotator cuff pathology, especially where timely MRI access is limited.

*For orthopaedic practice*, a reliable and accessible first-line imaging strategy may support earlier tear detection, referral, and treatment planning, while reserving MRI for equivocal or complex cases [27]. Advances in portable and high-resolution ultrasound further support its growing first-line role, while emerging AI-assisted approaches may improve diagnostic accuracy and reduce interobserver variability [28].

### Rationale for this review

This review focuses specifically on the supraspinatus tendon, the most commonly affected rotator cuff tendon and one that is readily assessed using ultrasound. Studies published from 2015 onward were included to reflect contemporary ultrasound technology and clinical practice. Earlier systematic reviews often incorporated older imaging technologies or evaluated the rotator cuff collectively, limiting tendon- and tear-specific interpretation [13, 16, 29]. More recent work has largely provided qualitative comparisons of US and MRI [30], leaving an important gap in contemporary quantitative evidence.

### The key contribution of the present review is a supraspinatus-specific, tear-stratified quantitative assessment of ultrasound performance

Rather than combining rotator cuff tears into a single diagnostic category, we separately evaluate any, partial-thickness, and full-thickness supraspinatus tears, allowing diagnostic performance to be interpreted according to tear severity. This distinction is clinically important because partial- and full-thickness tears differ in diagnostic difficulty and may influence management decisions. By integrating pooled diagnostic estimates with accuracy, predictive values, agreement measures, study quality, operator expertise, and equipment-related variability, this review provides a more clinically informative assessment of where ultrasound performs reliably and where MRI remains complementary. The review also identifies important evidence gaps, including the limited evaluation of handheld ultrasound and the absence of dedicated POCUS-based shoulder pathways.

## 2 Material and Methods

### 2.1 Search Strategy

This systematic review was conducted in accordance with the PRISMA (Preferred Reporting Items for Systematic Reviews and Meta-Analyses) guidelines [31]. A comprehensive literature search was conducted in MEDLINE, Embase, Scopus, and CINAHL Plus for English-language studies published between January 1, 2015, and June 1, 2026. The search strategy combined keywords, subject headings, and Boolean operators related to *ultrasound, magnetic resonance imaging, rotator cuff tears, supraspinatus, diagnostic accuracy*. Database-specific search strategies were adapted to the indexing and syntax requirements of each database and are provided in Table 1. Reference lists of included studies were also screened to identify additional eligible articles. Covidence was used for duplicate removal and study screening (Veritas Health Innovation, Melbourne, Australia; https://www.covidence.org/).

**Table 1.** Search strategies applied across the different databases.

| Database | Search Strategy |
| --- | --- |
| **CINAHL Plus with Full Text** | 1: (MH "Ultrasonography") OR (Ultrasound OR ultrasonograph*)<br>2: (MH "Magnetic Resonance Imaging") OR ("Gold standard" OR MRI)<br>3: (MH "Rotator Cuff") OR ((supraspinatus OR subscapularis OR infraspinatus OR "teres minor") N6 (tear* OR retear* OR torn OR rupture* OR lesion* OR injury*))<br>4: (MH "Sensitivity and Specificity") OR (sensitivity OR specificity OR "diagnostic accuracy" OR "reference standard" OR "predictive value*" OR PPV OR NPV OR "false positive*" OR "false negative*" OR "true positive*" OR "true negative*" OR "likelihood ratio*" OR "receiver operating characteristic*" OR "ROC curve*")<br>5: S1 AND S2 AND S3 AND S4 |
| **Embase and MEDLINE** | 1: ultrasound/ 263072; 2: (Ultrasound OR ultrasonograph*).tw,kf. 727131; 3: 1 OR 2 799195; 4: nuclear magnetic resonance imaging/ 1152064<br>5: ("Gold standard" OR MRI OR "magnetic resonance imaging").tw,kf. 1035630; 6: 4 OR 5 1514139<br>7: rotator cuff injury OR rotator cuff rupture/ 15751<br>8: (("rotator cuff" OR supraspinatus OR subscapularis OR infraspinatus OR "teres minor") (tear* OR retear* OR torn OR rupture* OR lesion* OR avulsion* OR injury*)).mp. 20675<br>9: 7 OR 8 20675; 10: accuracy 346741; 11: "sensitivity and specificity"/ 554449<br>12: (sensitivity OR specificity OR "diagnostic accuracy" OR "reference standard" OR "predictive value*" OR PPV OR NPV OR "false positive*" OR "false negative*" OR "true positive*" OR "true negative*" OR "likelihood ratio*" OR "receiver operating characteristic*" OR "ROC curve*").mp. 3146780<br>13: 10 OR 11 OR 12 3146780; 14: 3 AND 6 AND 9 AND 13 295<br>15: limit 14 to (conference abstracts OR "preprints (unpublished, non-peer reviewed)") 29<br>16: 14 NOT 15 266; 17: limit to yr = "2015–Current" 189 |
| **Scopus** | TITLE-ABS (Ultrasound OR ultrasonograph*) AND TITLE-ABS-KEY ("Gold standard" OR MRI OR "magnetic resonance imaging") AND TITLE-ABS-KEY (("rotator cuff" OR supraspinatus OR subscapularis OR infraspinatus OR "teres minor") W/6 (tear* OR retear* OR torn OR rupture* OR lesion* OR avulsion* OR injury*)) AND TITLE-ABS-KEY (sensitivity OR specificity OR "diagnostic accuracy" OR "reference standard" OR "predictive value*" OR PPV OR NPV OR "false positive*" OR "false negative*" OR "true positive*" OR "true negative*" OR "likelihood ratio*" OR "receiver operating characteristic*" OR "ROC curve*") AND PUBYEAR >= 2015 AND PUBYEAR <= 2026 |

### 2.2 Study Selection and Eligibility Criteria

Eligibility criteria were defined using the PICO framework [32]. The population included subjects with shoulder pain or suspected rotator cuff pathology, with a specific focus on supraspinatus tendon tears. Ultrasound was the index test, and MRI served as the noninvasive reference imaging standard. Outcomes of interest included sensitivity, specificity, accuracy, positive predictive value (PPV), and negative predictive value (NPV) for detecting supraspinatus tears.

Original, full-text clinical studies were eligible if they directly compared ultrasound with MRI and reported supraspinatus-specific quantitative diagnostic outcomes. Animal and cadaveric studies, conference abstracts, reviews, systematic reviews, meta-analyses, case reports, non-English publications, and studies without quantitative diagnostic data were excluded. Studies with fewer than 20 participants were also excluded to reduce the influence of very small samples on diagnostic estimates. For the quantitative meta-analysis, only studies for which complete 2 × 2 diagnostic data with true positives (TP), false negatives (FN), true negatives (TN), and false positives (FP) were reported or could be derived were included.

### 2.3 Data Extraction

Two authors (K.K. and S.G.) independently screened titles, abstracts, and full texts, with disagreements resolved through discussion and consensus or, when necessary, by a third author (A.H.). Data from included studies were extracted into a standardized Microsoft Excel template. K.K. performed the initial extraction, and S.G. independently verified the data for accuracy. Extracted variables included study design, sample size, population characteristics, ultrasound system, operator experience, tear type, reference standard, and diagnostic outcomes. These included sensitivity, specificity, agreement statistics (*κ* values and p-values), and study-level results for normal, partial-thickness, and full-thickness supraspinatus tears.

To enable formal meta-analysis, 2 × 2 diagnostic data comprising TP, FP, FN, and TN were extracted separately for any supraspinatus tear, partial-thickness supraspinatus tears, and full-thickness supraspinatus tears wherever available. Exact Clopper–Pearson 95% confidence intervals for sensitivity and specificity were also calculated for each tear category. Notes on study limitations and other relevant methodological considerations were recorded during data extraction.

### 2.4 Quality and Risk of Bias Assessment

The risk of bias and applicability of the included studies were evaluated using the Quality Assessment of Diagnostic Accuracy Studies–2 (QUADAS-2) tool [33]. Two authors (K.K. and S.G.) independently assessed each study across four domains for risk of bias: patient selection, index test, reference standard, and flow and timing, and across three domains for applicability: patient selection, index test, and reference standard. Each domain was rated as low, high, or unclear risk of bias or concern regarding applicability. In this review, ultrasound was considered the index test and MRI the reference imaging standard. Any disagreements between reviewers were resolved through discussion and consensus. Detailed findings of the QUADAS-2 assessment are presented in the Results section.

### 2.5 Diagnostic Meta-analysis

Due to variability in study design and reporting, complete 2 × 2 data were available for only a subset of studies. Where available or derivable, TP, FP, FN, and TN were used to assess ultrasound diagnostic accuracy against MRI. Studies lacking sufficient 2 × 2 data were retained in the qualitative synthesis but excluded from the quantitative meta-analysis.

Hierarchical Summary Receiver Operating Characteristic (HSROC) analyses and forest plots were generated using the metandi and midas packages in Stata version 18.0 [34, 35]. These analyses were used to obtain pooled estimates of sensitivity and specificity with corresponding 95% confidence intervals and to visualize variability in diagnostic performance across studies. Separate meta-analyses were conducted for any supraspinatus tear, partial-thickness supraspinatus tears, and full-thickness supraspinatus tears, allowing diagnostic performance to be evaluated according to tear severity.

## 3 Results

### 3.1 Study Selection

A total of 615 records were identified through database searches, including Embase (n = 229), Scopus (n = 198), MEDLINE (n = 140), and CINAHL (n = 48), and imported into Covidence. After removal of 331 duplicates, 284 records remained for title and abstract screening within the prespecified publication period of January 1, 2015, to June 1, 2026. Following screening against the predefined eligibility criteria, 86 full-text articles were assessed for eligibility. Of these, 63 were excluded because of a wrong comparator (n = 27), wrong study design (n = 14), wrong patient population (n = 8), irrelevant outcomes (n = 13), or inability to retrieve the full text (n = 1). Finally, 23 studies met the inclusion criteria and were included in the systematic review. The study selection process is illustrated in the PRISMA flow diagram Fig. 1. The 23 included studies formed the basis of the qualitative synthesis. Complete 2 × 2 diagnostic data comparing ultrasound with MRI were available for a subset of studies and were included in the quantitative meta-analysis. Specifically, 11 studies provided data for detection of any supraspinatus tear, 9 for full-thickness tears, and 8 for partial-thickness tears. Only 3 studies provided complete 2 × 2 data for all three diagnostic categories.

**Fig. 1.**
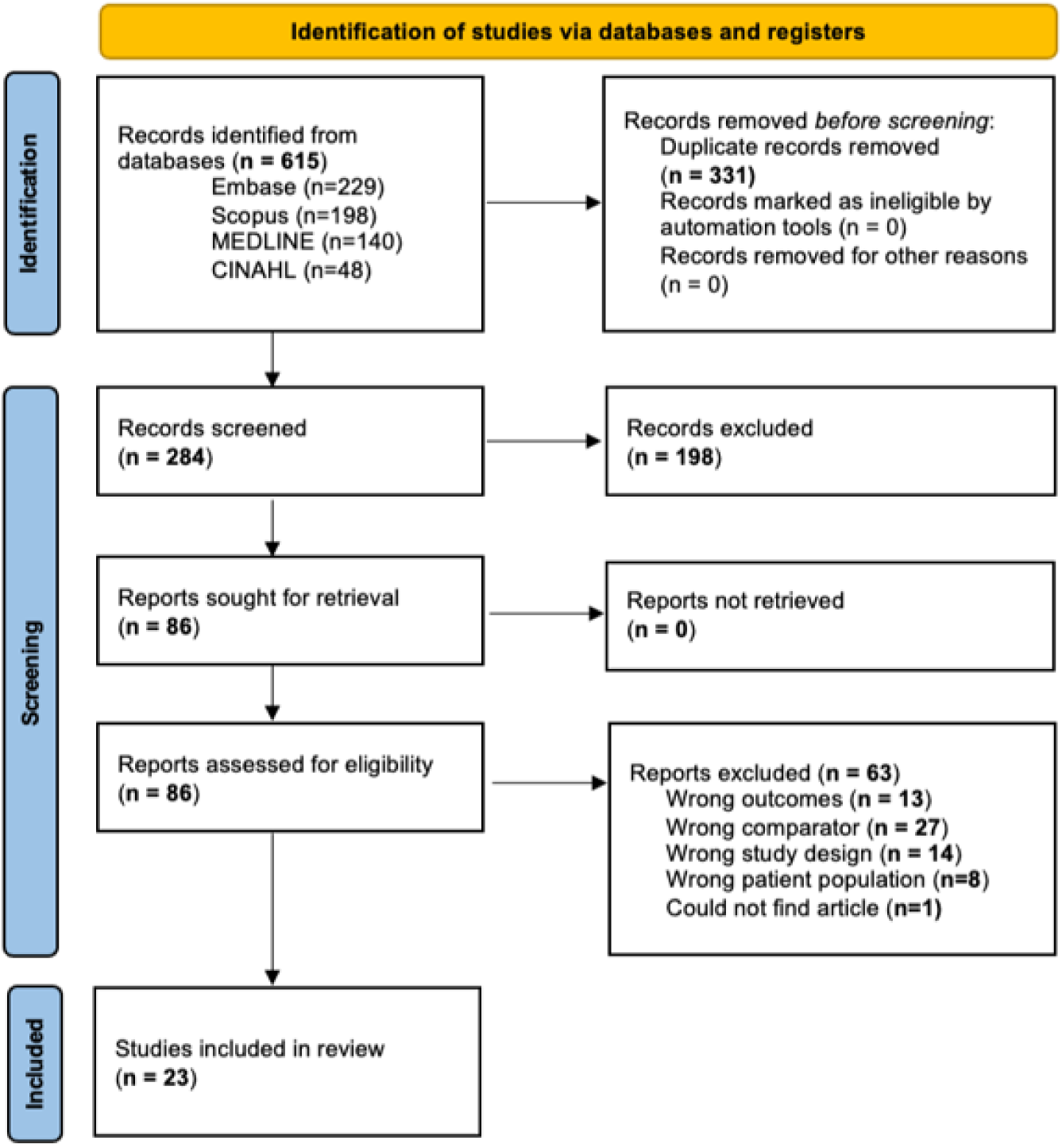
PRISMA flow diagram illustrating the identification, screening, eligibility assessment, and inclusion of studies in the systematic review.

### 3.2 Study Characteristics

The 23 included studies were published between 2015 and 2026 and reflected contemporary ultrasound technology and clinical practice. Of these, 17 were prospective, four retrospective, and two cross-sectional studies. Sample sizes ranged from 25 to 138 participants, comprising a total of 1,452 participants. Where reported, mean participant age ranged from 38.8 to 68.3 years. Most studies evaluated both partial- and full-thickness supraspinatus tears, while two studies focused on partial-thickness tears only. Most studies used conventional ultrasound systems with high-frequency linear-array transducers, with reported frequencies ranging from approximately 3 to 18 MHz. One study specifically evaluated a handheld ultrasound system.

Operators included radiologists, musculoskeletal radiologists, orthopaedic surgeons, sonographers, and a radiology trainee under specialist supervision, although operator experience was not reported in several studies. The characteristics of the included studies are summarized in Table 2.

**Table 2.** Characteristics of the included studies comparing supraspinatus US with MRI (n = 23).

| Author (Year) | Study Design | Sample size (SS) and Mean age (MA) | Tear | US scanner | US operator |
| --- | --- | --- | --- | --- | --- |
| **Banerjee (2025) [50]** | Prospective | SS = 50;<br>MA = NS | Partial and Full | GE Voluson S6 Pro (7-12MHz) | Radiologist |
| **Barad (2022) [55]** | Prospective | SS = 50;<br>MA = 51.2 | Partial | NS | NS |
| **Elgamal (2026) [40]** | Prospective | SS = 25;<br>MA = 46.4 | Partial and Full | Toshiba Ultrasound Aplio 500 | Two radiologists (14 and 7 years) of MSK experience |
| **Fischer (2015) [45]** | Prospective | SS = 45;<br>MA = 68.3 | Partial and Full | GE Logiq P5 (10 MHz) | Orthopedic surgeon with over ten years of experience in shoulder US |
| **Furtado (2024) [51]** | Prospective | SS = 112;<br>MA = NS | Partial | Siemens ACUSON S2000 (7.5-14MHz) | Radiologist |
| **Ganesh (2024) [49]** | Prospective | SS = 53;<br>MA = 48.6 | Partial and Full | GE Voluson S8 (9 MHz) | Radiologist specializing in MSK imaging with eight years of experience |
| **Ghosh (2026) [46]** | Retrospective | SS = 138;<br>MA = 51.5 | Partial and Full | GE LogicE9 and ML6-15-D linear array transducers | NS |
| **Gupta (2022) [41]** | Prospective | SS = 50;<br>MA = 56 | Partial and Full | GE Voluson E6 / Samsung HS70A (8-15 MHz) | Radiology resident under supervision of a Senior Radiologist |
| **Khan (2024) [47]** | Prospective | SS = 40;<br>MA = NS | Partial and Full | Samsung HS70A | Radiologist |
| **Kraats (2023) [36]** | Retrospective | SS = 61;<br>MA = 64 | Partial and Full | Philips Lumify L12-4 linear array transducer | Orthopedic surgeon and MSK radiologist |
| **Mandloi (2024) [56]** | Prospective | SS = 50;<br>MA = NS | Partial and Full | NS | NS |
| **Mohtasib (2019) [37]** | Retrospective | SS = 86;<br>MA = 53.7 | Partial and Full | Philips IU22, EPICQ7 (3-12 MHz) | MSK Radiologist and Radiologist fellow |
| **Murugan (2023) [52]** | Prospective | SS = 90;<br>MA = 40.5 | Partial and Full | Philips Affiniti 70G (12-3 MHz) | Radiologist |
| **Nunna (2024) [54]** | Prospective | SS = 80;<br>MA = NS | Partial and Full | Aloka Hitachi (12-18 MHz) | NS |
| **Padhy (2023) [57]** | Prospective | SS = 30;<br>MA = 52 | Partial and Full | NS | NS |
| **Pandya (2025) [42]** | Prospective | SS = 84;<br>MA = NS | Partial and Full | Conventional USG | NS |
| **Paul (2026) [58]** | Cross-sectional | SS = 70;<br>MA = 39.7 | Partial and Full | NS | NS |
| **Rajkumar (2025) [43]** | Prospective | SS = 45;<br>MA = 38.8 | Partial and Full | Philips HD 11 XE (12-3 MHz) | Radiologist |
| **Roshini (2024) [44]** | Prospective | SS = 40;<br>MA = 43 | Partial and Full | Mindray DC8 (7-12 MHz) | NS |
| **Stephen (2026) [59]** | Cross-sectional | SS = 60;<br>MA = 67.6 | Partial and Full | Mindray DC 80 (6-12 MHz) | NS |
| **Varma (2024) [38]** | Prospective | SS = 35;<br>MA = 7NS | Partial and Full | Philips Affinity 70 (5-12 MHz) | MSK Sonographer |
| **Venkatesh (2025) [39]** | Retrospective | SS = 78;<br>MA = 43.7 | Partial and Full | Samsung HS70A, GE Voluson S8, Samsung HS40 | NS |
| **Yazigi (2018) [53]** | Prospective | SS = 80;<br>MA = 48.9 | Partial and Full | NR (10 MHz) | Two Radiologists |

NS = Not Specified; NR = Not Reported

### 3.3 Risk of Bias Assessment Results

Overall, most studies demonstrated a low risk of bias in the patient-selection domain, although Kraats et al. [36], Mohtasib et al. [37], Varma et al. [38], and Venkatesh et al. [39] were judged to be at high risk. In the index-test and reference-standard domains, several studies were rated as having some concerns, largely because of insufficient reporting of methodological safeguards such as blinding. Elgamal et al. [40] was the only study judged to be at high risk in the index-test domain, while Mohtasib et al. [37] was the only study rated at high risk in the reference-standard domain. Flow and timing also frequently raised some concerns, with Kraats et al. [36] and Mohtasib et al. [37] being the only two studies judged to be at high risk in this domain (see Fig. 2 for details).

**Fig. 2.**
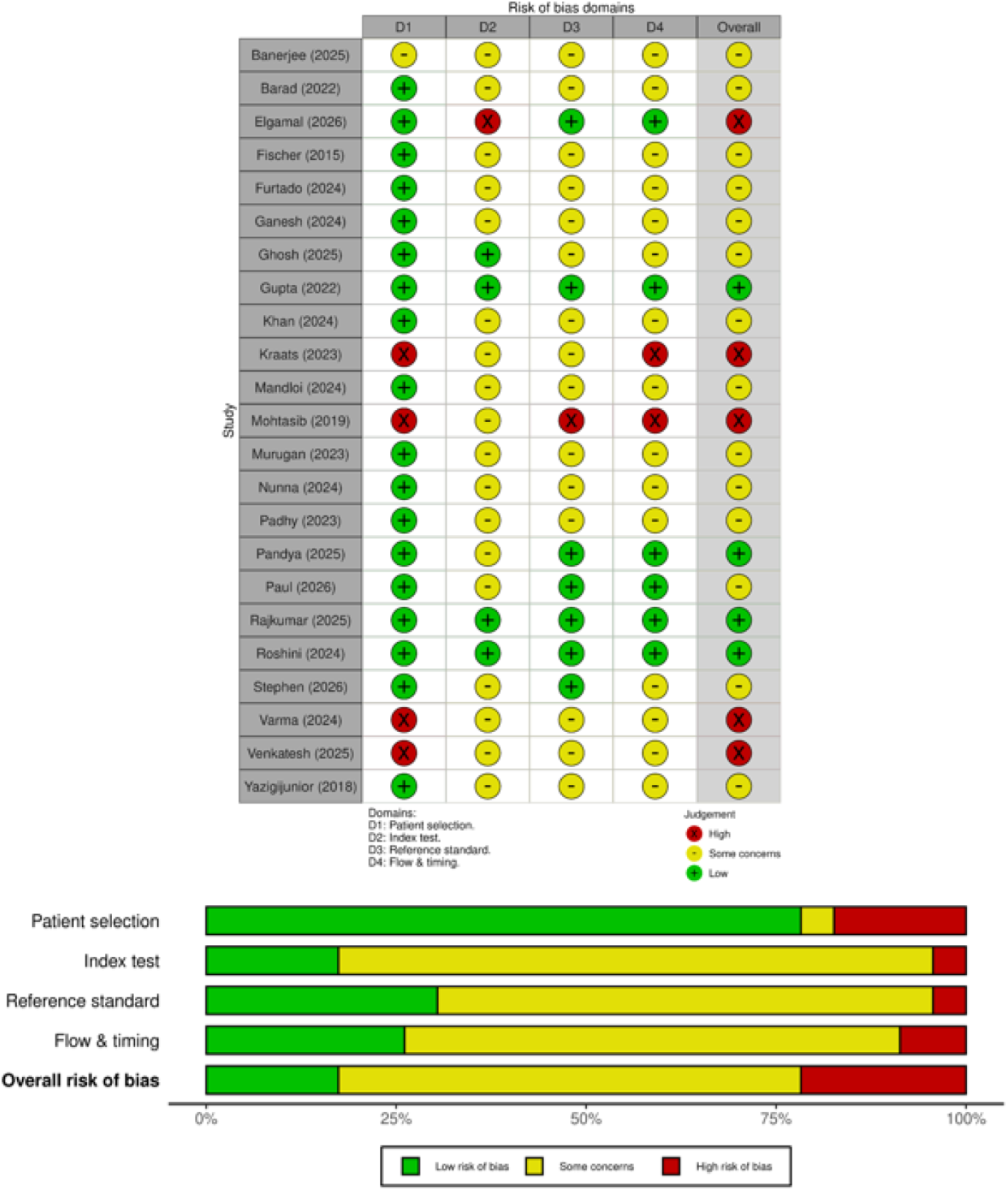
QUADAS-2 quality assessment of the included diagnostic accuracy studies. The upper panel shows study-level risk-of-bias judgments across the four QUADAS-2 domains and overall assessment, while the lower panel summarizes the proportion of studies rated as low risk, some concerns, or high risk of bias.

Overall risk-of-bias judgments were predominantly characterized by some concerns. Five studies were judged to have an overall high risk of bias: Elgamal et al. [40], Kraats et al. [36], Mohtasib et al. [37], Varma et al. [38], and Venkatesh et al. [39]. Four studies—Gupta et al. [41], Pandya et al. [42], Rajkumar et al. [43], and Roshini et al. [44] were judged to have an overall low risk of bias, while the remaining 14 studies were rated as having some concerns. These concerns were most commonly related to insufficient reporting or methodological limitations involving the index test, reference standard, and flow-and-timing domains.

### 3.4 Diagnostic Accuracy

#### 3.4.1 Overall Diagnostic Performance

Diagnostic performance was assessed against MRI, with tear-specific sensitivity, specificity, PPV, NPV, and accuracy summarized in Table 3. Studies reporting combined tear outcomes were summarized separately in Table 4 (A). Where complete 2 × 2 data were available, studies were also included in the meta-analysis of any supraspinatus tear.

**Table 3.**
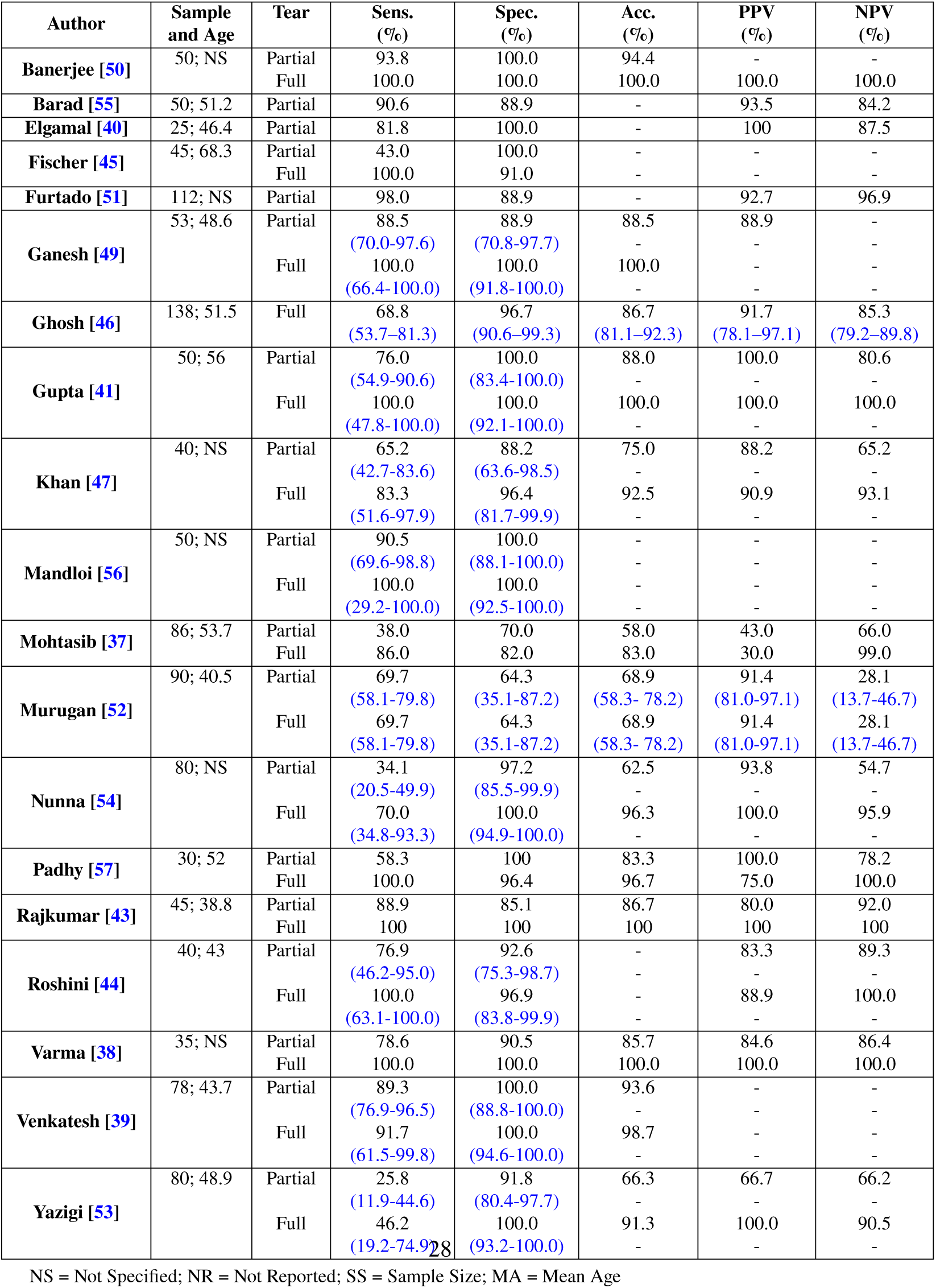
Comparison of ultrasound diagnostic performance for partial- and full-thickness supraspinatus tears assessed separately, using MRI as the reference standard (sensitivity, specificity, PPV, NPV, and overall diagnostic accuracy).

**Table 4.** Diagnostic performance and agreement statistics for ultrasound assessment of supraspinatus tears using MRI as the reference standard. (A). Combined partial- and fullthickness diagnostic outcomes. (B). Reported kappa (*κ*) statistics and p-values.

| **A. Combined partial- and full-thickness diagnostic outcomes** |  |  |  |  |  |  |  |
| --- | --- | --- | --- | --- | --- | --- | --- |
| **Author** | **Sample and Age** | **Tear** | **Sens. (%)** | **Spec. (%)** | **Acc. (%)** | **PPV (%)** | **NPV (%)** |
| **Kraats [36]** | 61; 64 | Combined | 81.1 | 62.5 | - | - | - |
| **Pandya [42]** | 84; NS | Combined | 83.8 | 52.1 | 79.9 | 91.4 | 26.4 |
| **Paul [58]** | 70; 39.7 | Combined | 84.0 | 78.8 | - | - | - |
| **Stephen [59]** | 60; 67.6 | Combined | 88.3 | - | - | 100 | - |

| **B. Reported kappa ($\kappa$) statistics and p-values** |  |  |  |  |
| --- | --- | --- | --- | --- |
| **Barad [55]** | SS = 50; | Partial | 0.75 | - |
|  | MA = 51.2 | Full | - | - |
| **Fischer [45]** | SS = 45; | Partial | 0.85 | 0.68 |
|  | MA = 68.3 | Full | 0.85 | 0.26 |
| **Furtado [51]** | SS = 112; | Partial | 0.88 | < 0.0001 |
|  | MA = NS | Full | - | - |
| **Ganesh [49]** | SS = 53; | Partial | - | 0.0012 |
|  | MA = 48.6 | Full | - | 0.0021 |
| **Ghosh [46]** | SS = 138; | Partial | 0.59<br>(0.46–0.72) | < 0.0001 |
|  | MA = 51.5 | Full | 0.53<br>(0.34–0.70) | < 0.0001 |
| **Gupta [41]** | SS = 50; | Partial | 0.76 | - |
|  | MA = 56 | Full | 1.0 | - |
| **Khan [47]** | SS = 40; | Partial | 0.51 | - |
|  | MA = NS | Full | 0.82 | - |
| **Padhy [57]** | SS = 30; | Partial | - | 0.5 |
|  | MA = 52 | Full | - | 0.4 |
| **Varma [38]** | SS = 35; | Partial | - | < 0.001 |
|  | MA = NS | Full | - | - |
| **Venkatesh [39]** | SS = 78; | Partial | 0.88 | < 0.0001 |
|  | MA = 43.7 | Full | 0.94 | < 0.0001 |
| **Yazigi [53]** | SS = 80; | Partial | - | 0.031 |
|  | MA = 48.9 | Full | - | < 0.001 |

NS = Not Specified; SS = Sample Size; MA = Mean Age; Sens. = Sensitivity; Spec. = Specificity; Acc. = Accuracy; PPV = Positive Predictive Value; NPV = Negative Predictive Value.

Across studies with tear-specific data, ultrasound demonstrated stronger and more consistent diagnostic performance for full-thickness than for partial-thickness supraspinatus tears. For partial-thickness tears, mean sensitivity, specificity, and accuracy were 71.5% ± 22.6%, 91.5% ± 10.3%, and 79.2% ± 12.6%, respectively. For full-thickness tears, the corresponding values were 88.5% ± 16.5%, 95.2% ± 9.5%, and 93.4% ± 9.2%, respectively. These descriptive study-level averages indicate higher diagnostic performance and lower variability for full-thickness supraspinatus tears, particularly for sensitivity and overall accuracy.

#### 3.4.2 Partial-thickness supraspinatus tears

Across the 18 studies reporting separate diagnostic performance for partial-thickness supraspinatus tears, sensitivity ranged from 25.8% to 98.0%. Eleven studies (61.1%) reported sensitivity ≥ 70%, while seven (38.9%) reported sensitivity < 70%. Specificity ranged from 64.3% to 100%, with 11 of 18 studies (61.1%) reporting specificity ≥ 90%. Overall accuracy, where reported, ranged from 58.0% to 94.4%.

#### 3.4.3 Full-thickness supraspinatus tears

Across the 16 studies reporting separate diagnostic performance for full-thickness supraspinatus tears, sensitivity ranged from 46.2% to 100%, with 10 studies (62.5%) reporting sensitivity ≥ 90%. Specificity ranged from 64.3% to 100%, with 14 of 16 studies (87.5%) reporting specificity ≥ 90%. Where reported, PPV ranged from 30.0% to 100% and NPV from 28.1% to 100%. Overall accuracy ranged from 68.9% to 100%, with 10 of 13 studies (76.9%) reporting accuracy ≥ 90%. Overall, diagnostic performance was higher and more consistent for fullthickness than for partial-thickness supraspinatus tears. Study-level variation in sensitivity, specificity, and diagnostic accuracy for partial- and full-thickness tears is illustrated in Fig. 3.

**Fig. 3.**
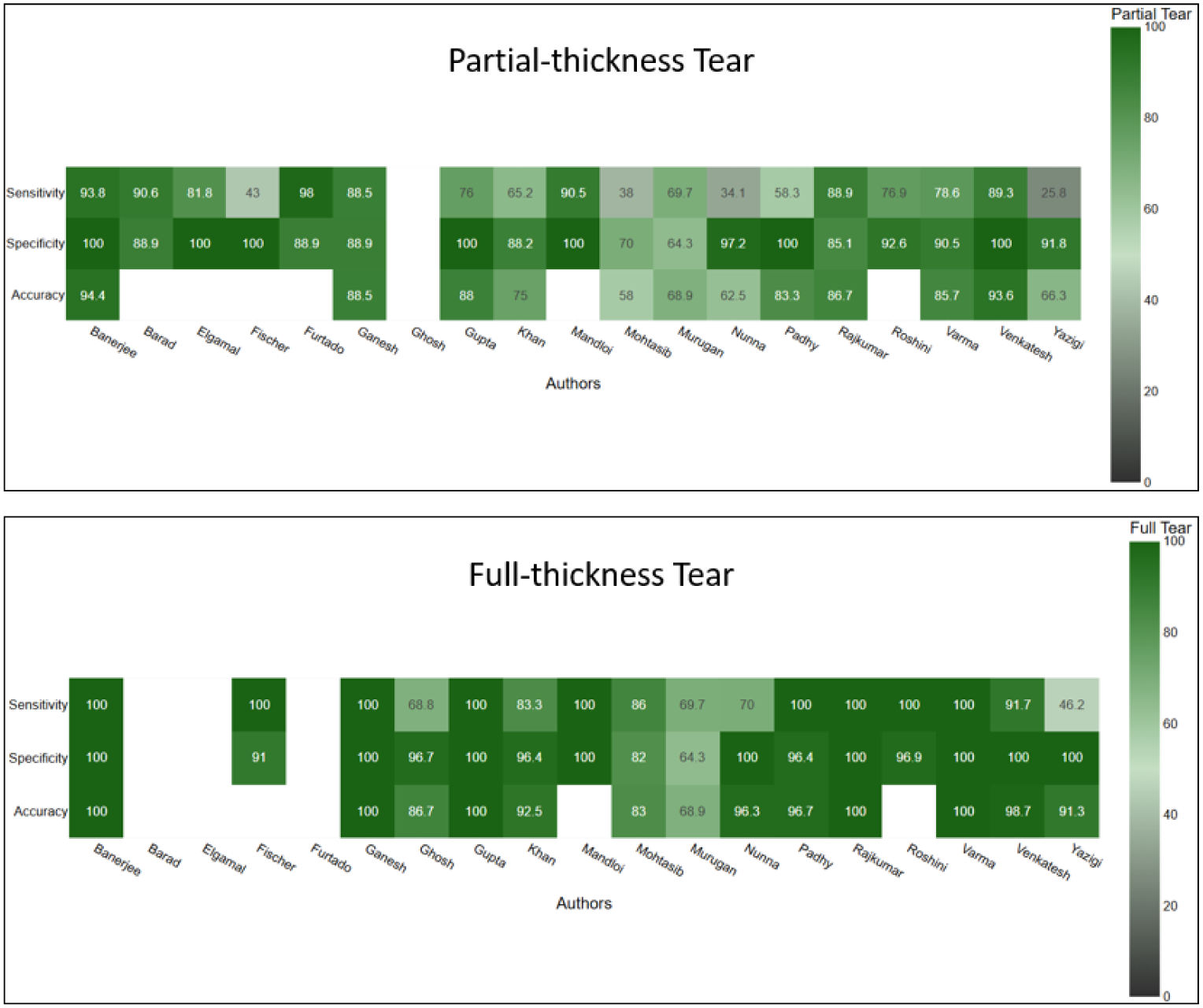
Heat maps showing sensitivity, specificity, and diagnostic accuracy for partial-thickness supraspinatus tears (top) and full-thickness supraspinatus tears (bottom). Blank cells indicate studies that did not report the corresponding diagnostic metric.

### 3.5 Diagnostic Meta-analysis

Formal diagnostic meta-analysis was performed using MRI as the reference imaging standard. Separate analyses were conducted for any supraspinatus tear, partial-thickness supraspinatus tears, and full-thickness supraspinatus tears. HSROC analyses and forest plots were used to summarize pooled sensitivity and specificity and their corresponding 95% confidence intervals.

#### 3.5.1 Any supraspinatus tear

HSROC analysis of the 11 studies with complete 2 × 2 table data on detection of any tear available showed an overall sensitivity and specificity of 96% (95% CI: 86-98%) and 76% (95% CI: 58-87%), respectively, as shown in Fig. 4 (A).

**Fig. 4.**
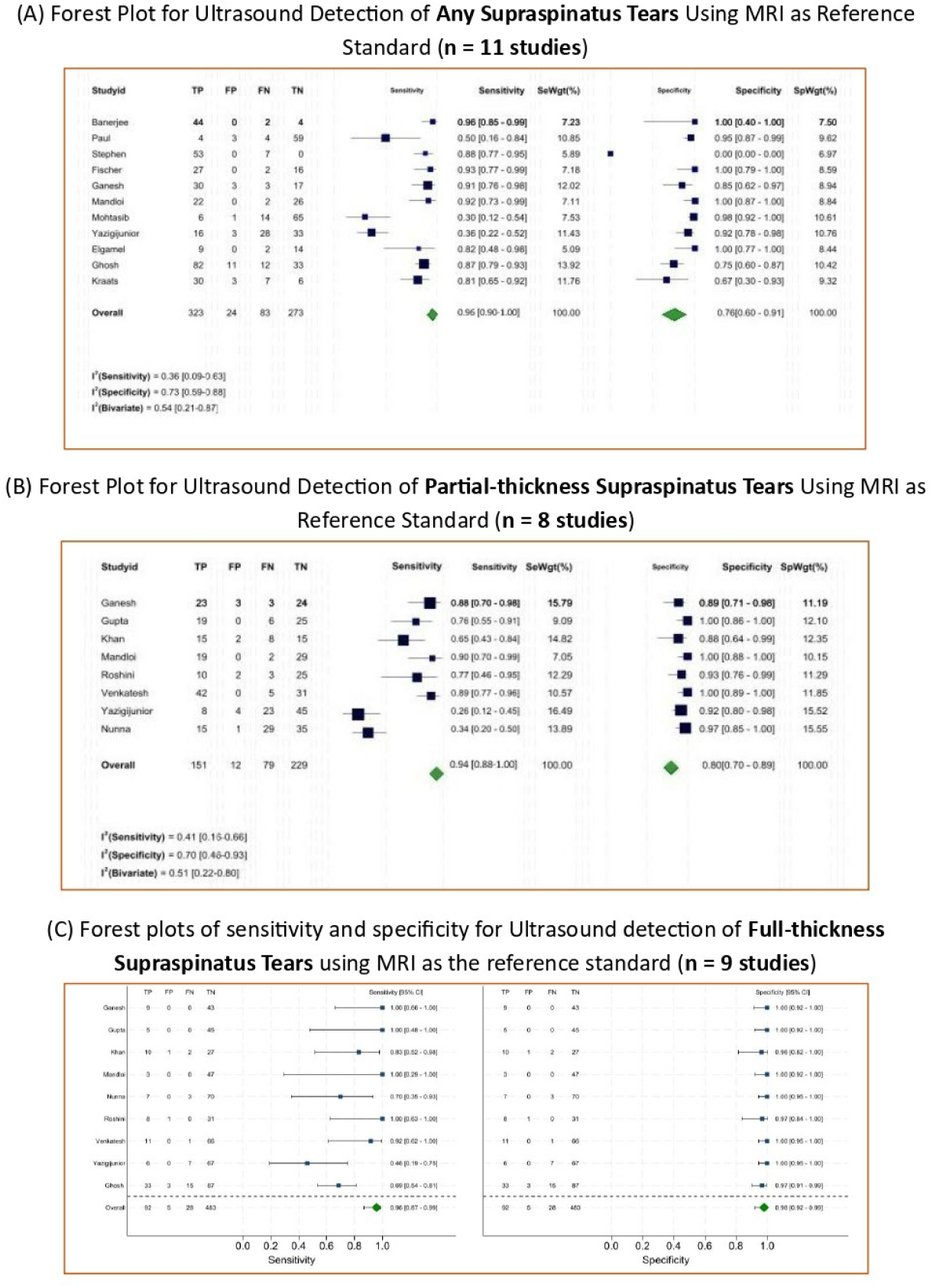
Forest plots of sensitivity and specificity for ultrasound detection of supraspinatus tears using MRI as the reference imaging standard: (A) any supraspinatus tear (n = 11 studies), (B) partial-thickness tears (n = 8 studies), and (C) full-thickness tears (n = 9 studies).

#### 3.5.2 Partial-thickness supraspinatus tear

Eight studies provided complete 2 × 2 diagnostic data for partial-thickness supraspinatus tears. HSROC analysis showed a pooled sensitivity of 94% (95% CI: 87–100%) and a pooled specificity of 80% (95% CI: 70–89%), as presented in Fig. 4 (B). The wider variation observed among individual studies was consistent with the greater heterogeneity in ultrasound performance for partial-thickness tears identified in the descriptive analysis.

#### 3.5.3 Full-thickness supraspinatus tear

Nine studies provided complete 2 × 2 diagnostic data for full-thickness supraspinatus tears. HSROC analysis demonstrated a pooled sensitivity of 96% (95% CI: 87–99%) and a pooled specificity of 98% (95% CI: 92-99%), (see Fig. 4 (C)). Overall, the quantitative findings indicated strong diagnostic performance of ultrasound for full-thickness supraspinatus tears.

### 3.6 Kappa Values and Statistical Significance

Agreement was reported using kappa (*κ*) statistics in several studies. Five studies (Fischer, Ghosh, Gupta, Khan, and Venkatesh) [39, 41, 45–47] reported *κ* values for both partial- and full-thickness supraspinatus tears. Among these, Fischer, Ghosh, and Venkatesh also reported associated p-values. Across seven studies reporting *κ* values for partial-thickness tears, values ranged from 0.51 to 0.88. For full-thickness tears, five studies reported *κ* values ranging from 0.53 to 1.00. Overall, agreement tended to be higher and more consistent for full-thickness tears, although the type of agreement assessed and reporting methods varied across studies. Several additional studies reported p-values without corresponding *κ* estimates, limiting direct comparison of agreement across the full evidence base. Reported *κ* values and associated p-values are summarized in Table 4, (B).

## 4 Discussion

This systematic review and meta-analysis provides a contemporary evaluation of ultrasound for supraspinatus tendon tears using MRI as the non-invasive reference imaging standard. Across 23 studies involving 1,452 participants, ultrasound demonstrated stronger and more consistent performance for full-thickness than for partial-thickness tears. Mean sensitivity, specificity, and accuracy were higher for full-thickness tears (88.5%, 95.2%, and 93.4%) than for partial-thickness tears (71.5%, 91.5%, and 79.2%), respectively. This tear-specific difference is clinically important and highlights the need to interpret ultrasound performance according to tear severity.

Full-thickness tears produce complete structural disruption, whereas partial-thickness tears may be small, articular- or bursal-sided, and difficult to distinguish from tendinosis or imaging artefacts. Ultrasound is also susceptible to anisotropy, whereby subtle changes in probe orientation alter tendon echogenicity and may obscure small defects [48]. These factors likely explain the lower and more variable sensitivity observed for partial-thickness tears despite generally high specificity. This pattern suggests that ultrasound is more consistent for confirming obvious structural abnormalities than for excluding subtle partial-thickness pathology. Clinically, a negative or equivocal ultrasound examination should therefore be interpreted cautiously when suspicion for a partial-thickness tear remains high, supporting a complementary role for MRI in such cases.

The present findings are consistent with earlier evidence showing stronger ultrasound performance for full-thickness than partial-thickness tears. De Jesus et al. [13] found no significant difference between US and MRI, although their analysis included older imaging technologies and a broader rotator cuff population. Smith et al. [29] reported higher pooled sensitivity and specificity for full-thickness than partial-thickness tears, and Farooqi et al. [16] similarly found lower ultrasound performance for partial-thickness supraspinatus tears. Together, these studies support the consistent observation that full-thickness tears are detected more reliably. Our review extends this evidence by focusing specifically on the supraspinatus tendon and contemporary studies from 2015 onward. Compared with the recent 2026 review [30], which included 13 studies and was primarily qualitative, our review includes 23 studies and provides separate quantitative analyses for any, partial-thickness, and full-thickness tears. This tear-stratified approach reveals clinically important differences in diagnostic performance that may be obscured when tear categories are combined and provides a more clinically informative assessment of the role of ultrasound across tear severity.

Operator expertise and ultrasound equipment varied considerably across studies. Some involved experienced MSK radiologists or orthopaedic surgeons with more than eight years of shoulder ultrasound experience [45, 49], whereas others used radiologists, supervised trainees, or sonographers [38, 41, 47, 50–53]; several studies did not report operator experience [39, 44, 54–57]. Most studies used conventional high-frequency systems, while only van der Kraats et al. [36] evaluated a handheld device. That study reported 81.1% sensitivity and 62.5% specificity, with high intrarater but lower interrater agreement, highlighting persistent operator dependence. Overall, these findings emphasize the importance of operator training, standardized scanning protocols, and further validation of handheld and POCUS systems, particularly for partial-thickness tears.

Studies reporting partial- and full-thickness tears as a combined group showed similar sensitivity but more variable specificity. Van der Kraats et al. [36], Pandya et al. [42], and Paul et al. [58] reported sensitivities of 81.1%, 83.9%, and 84.0%, respectively, while specificity ranged from 52.1% to 78.8%; Stephen et al. [59] reported a sensitivity of 88.3%, but specificity could not be estimated. These combined estimates are difficult to interpret because partial- and full-thickness tears differ substantially in diagnostic difficulty. Pooling them may therefore obscure the source of false-negative and false-positive findings and mask clinically important differences in ultrasound performance. This supports tear-stratified analysis whenever sufficient data are available.

Across studies reporting kappa (*κ*) statistics, agreement values ranged from 0.51 to 0.88 for partial-thickness tears and from 0.53 to 1.00 for full-thickness tears. High agreement for full-thickness tears was reported by Gupta et al. [41] (*κ* = 1.00) and Venkatesh et al. [39] (*κ* = 0.94), while agreement for partial-thickness tears was more variable. However, the type of agreement assessed and reporting methods differed across studies, limiting direct comparison of *κ* values. Several studies also reported p-values without corresponding *κ* estimates. Overall, the available evidence suggests more consistent agreement for full-thickness tears, although agreement reporting remained limited and heterogeneous.

The QUADAS-2 assessment showed mixed study quality, with four studies at low overall risk of bias, five at high risk, and the remaining 14 studies showing some concerns. High overall risk was identified in Elgamal et al. [40], van der Kraats et al. [36], Mohtasib et al. [37], Varma et al. [38], and Venkatesh et al. [39]. High-risk judgments were mainly related to patient selection, while concerns in the index test, reference standard, and flow/timing domains often reflected incomplete reporting of safeguards such as blinding. Delays between ultrasound and MRI may also have influenced diagnostic agreement by allowing changes in pathology. These methodological differences may partly explain variability in diagnostic estimates and should be considered when interpreting the pooled findings.

From an orthopaedic perspective, ultrasound and MRI have complementary roles in rotator cuff assessment. Ultrasound is an accessible, dynamic, and lower-cost first-line option, particularly for suspected full-thickness supraspinatus tears and in settings with limited MRI access [22, 27]. Its greater variability for partial-thickness tears supports continued use of MRI for subtle or equivocal lesions and for detailed assessment of tear extent, retraction, muscle changes, associated pathology, and preoperative planning [15, 27, 60]. Thus, ultrasound may reduce reliance on MRI in straightforward cases while MRI remains important for complex or uncertain findings.

A major strength of this review is the tear-stratified diagnostic meta-analysis, which assessed ultrasound performance separately for any, partial-thickness, and full-thickness supraspinatus tears. Eleven, eight, and nine studies contributed complete 2 × 2 data, respectively. Pooled sensitivity/specificity were 96% / 76% for any tear, 94% / 80% for partial-thickness tears, and 96% / 98% for full-thickness tears. The stronger performance for full-thickness tears was consistent with the descriptive findings. This stratified approach provides a more clinically interpretable assessment across tear severity, although pooled estimates were based on only subsets of the included studies.

Several limitations should be considered. Complete 2 × 2 data were available only for subsets of studies, limiting each meta-analysis. Heterogeneity in patient populations, ultrasound systems, operator expertise, scanning protocols, and reporting methods may also have influenced diagnostic estimates. Several studies had modest sample sizes, evidence for handheld ultrasound was limited to one study, and none evaluated a dedicated POCUS shoulder pathway. These factors may limit the generalizability of the pooled findings.

None of the included studies evaluated a dedicated POCUS shoulder pathway, despite its growing use in bedside and primary-care settings [25, 26]. Future studies should assess handheld and POCUS-based shoulder ultrasound using standardized protocols, defined operator training, and tear-specific reporting. AI-assisted approaches may also help reduce operator dependence and support less-experienced users [28]. Prospective multicentre studies should further evaluate diagnostic performance, clinical outcomes, cost-effectiveness, and downstream MRI use to clarify the role of ultrasound in real-world orthopaedic pathways.

## 5 Conclusion

Ultrasound demonstrated strong diagnostic performance for supraspinatus tears, particularly full-thickness tears, while partial-thickness tears remained more variable and challenging. Tear-stratified analysis provides greater clinical value than combining tear categories and supports ultrasound as an accessible first-line imaging modality for rotator cuff assessment. MRI remains complementary for equivocal or subtle partial-thickness tears, associated pathology, and preoperative planning. Standardized protocols, operator training, and continued evaluation of handheld and POCUS-based approaches may further strengthen the role of ultrasound in orthopaedic practice.

## Data Availability

All data analyzed in this systematic review and meta-analysis were obtained from previously published studies and are summarized within the manuscript and its tables. No new individual-level dataset was generated for this study.

